# Pre-extracorporeal support impaired oxygen delivery as a predictor of neurologic outcomes in infants; insights into decision support

**DOI:** 10.1101/2025.04.26.25320858

**Authors:** Edon J Rabinowitz, Amy Ouyang, Rejean Guerriero, Sarah Bauer Huang, Kristin Guilliams, Philip RO Payne, Ahmed S Said

## Abstract

**Objectives:** This study aims to evaluate the association between pre-Extracorporeal Membrane Oxygenation (ECMO) markers of impaired oxygen delivery, as quantified by the iDO2 predictive analytics platform, and neurologic outcomes in infants supported on ECMO. The goal is to determine whether these markers can inform decision-support systems for optimizing ECMO initiation timing.

**Materials & Methods:** We performed a single-center retrospective cohort study of infants <1 year supported on ECMO from 2013–2017, excluding cases with congenital diaphragmatic hernia or post-cardiac surgery ECMO. Data included demographics, clinical variables, and iDO2 estimates retroactively calculated in 120-minute intervals prior to ECMO initiation. Primary outcomes included mortality, EEG abnormalities, and head imaging findings; secondary outcomes included MRI abnormalities and Functional Status Scores (FSS).

**Results:** Of 219 patients, 47 met inclusion criteria. Median age and weight at ECMO initiation were 16 days [IQR 6–112] and 3.3 kg [IQR 2.8–4.8], with an overall mortality rate of 55%. Non-survivors had higher rates of congenital heart disease (77% vs. 42%, p=0.03) and pre-ECMO cardiac arrest (53% vs. 14%, p=0.006). Time spent above iDO2 thresholds of 25%, 50%, and 75% increased closer to ECMO initiation. Higher iDO2 dose correlated with adverse neurologic outcomes, including EEG abnormalities and abnormal imaging, and predicted poor composite functional outcomes (p<0.05).

**Discussion & Conclusion:** Markers of impaired oxygen delivery, such as iDO2, may inform the development of decision-support systems to optimize ECMO timing, potentially improving neurologic outcomes. Further research is needed to validate these findings and develop decision-support systems for clinical practice.

## BACKGROUND AND SIGNIFICANCE

Extracorporeal Membrane Oxygenation (ECMO) represents a high-risk and resource-intensive modality of support, requiring careful consideration in its application[1,2]. Decision-making for ECMO involves balancing the risks associated with the underlying pathology and subsequent impaired tissue oxygenation against the inherent risks of ECMO itself, particularly its potential neurologic complications. Despite the growing use of ECMO[3], the current literature reveals a significant gap in tools designed to support clinical decision-making for its initiation.

Existing tools predominantly focus on mortality prognostication, failing to account for the progression of physiological extremis, even though the degree of end-organ dysfunction is well established as a predictor of poor outcomes. In contrast, the intensive care unit (ICU) is now equipped with advanced tools that can estimate impaired oxygen delivery in near real time, such as the iDO2 platform. These tools provide insights beyond conventional metrics, offering the potential for enhanced clinical guidance.

Our study aimed to assess the association between pre-ECMO markers of impaired oxygen delivery and neurologic outcomes, extending the focus beyond mortality alone. Ultimately, we sought to determine whether these tools could inform the development of clinically applicable decision-support systems, enabling the optimal timing of ECMO initiation by leveraging evolving patient trajectories and markers of impaired oxygen delivery.

## MATERIALS AND METHODS

### Data Sources and Study Participants

We conducted a single center, retrospective cohort study of infants supported on ECMO from January 2013 – September 2017. The study was approved by the Washington University in St Louis Institutional Review Board with a waiver of informed consent (IRB # 201812081, approved December 16^th^, 2018).

We included infants, <1 year of age, supported on ECMO in the pediatric or cardiac ICU. This age group was chosen as all infants in this category would have at least one mode of head imaging performed during ECMO (e.g., head ultrasound) to serve as an outcome measure. Exclusion criteria included children >1 year of age, children with congenital diaphragmatic hernia (CDH) as the probability of utilizing ECMO support in this population is often known prenatally depending on the severity of CDH, and those placed on ECMO directly following cardiac surgery due to inability to separate from CPB or at a referring institution prior to transfer to our center.

### Predictor Variables

Review of the electronic health record (EHR) data was performed to collect patient and ECMO related variables including: demographics, gestational age, underlying pathology prior to ECMO initiation, presence and form of CHD and related surgical intervention, comorbidities, genetic abnormalities, mechanical ventilation, use of vasoactive medications and markers of oxygen delivery (e.g. urine output, near infrared spectroscopy; NIRS, biochemical markers of perfusion; lactate or mixed venous saturation). ECMO variables included duration of ECMO, mode of cannulation and circumstances of deployment (emergent vs elective), history of cardiac arrest prior to ECMO deployment and history of extracorporeal cardiopulmonary resuscitation (ECPR).

### High resolution marker of impaired oxygen delivery – iDO2

T3 Data Aggregation and Visualization (T3) is a predictive analytics application developed in 2010 by Etiometry LLC (Brighton, MA)[4]. It has been utilized in multiple ICUs across the United States over the past several years. T3 enables the collection, visualization, and storage of near real-time physiological data streams. Leveraging physiology-based models, it employs predictive analytics to estimate the probability of a state of inadequate oxygen delivery (iDO2).

The algorithm uses up to 13 live physiologic signals to estimate the probability (from 0-100) of a mixed venous saturation being <40%; the threshold below which anaerobic metabolism is triggered. iDO2 is continuously estimated in 5 second intervals and reflected in the T3 platform[5]. A minimum of 9 parameters and blood pressure recordings at 10-minute intervals are required to generate an iDO2 estimation. iDO2 has been validated against traditional markers of end organ oxygenation and FDA 510(K) cleared[6].

iDO2 data was calculated retroactively by incorporating continuous telemetry data directly extracted from the patients’ monitors and locally stored on a BedMasterEX ® server and relevant laboratory data and medical history from the EHR. This retroactive approach for data (prior to institutional deployment of the T3 platform) assured that study population or investigators were not exposed to T3 in real time and removes the possibility that iDO2 calculations influenced clinician decision making. iDO2 data was examined at 120-minute time intervals starting 6 hours prior to ECMO initiation time, 6 to 4 hours, 4 to 2 hours and 2 to zero hours prior to ECMO. For each 120-minute interval, the percentage of time spent over iDO2 probability thresholds of 25%, 50% and 75% were recorded (i.e. the higher the threshold and the longer the time, the worst the oxygen delivery state).

### iDO2 dose

To quantify the burden of iDO2, the weighted average of all iDO2 calculations (or the area under the IDO2 curve) in each 120-minute time interval was used to generate an iDO2 dose (**Figure 1a**).

**Figure 1:**
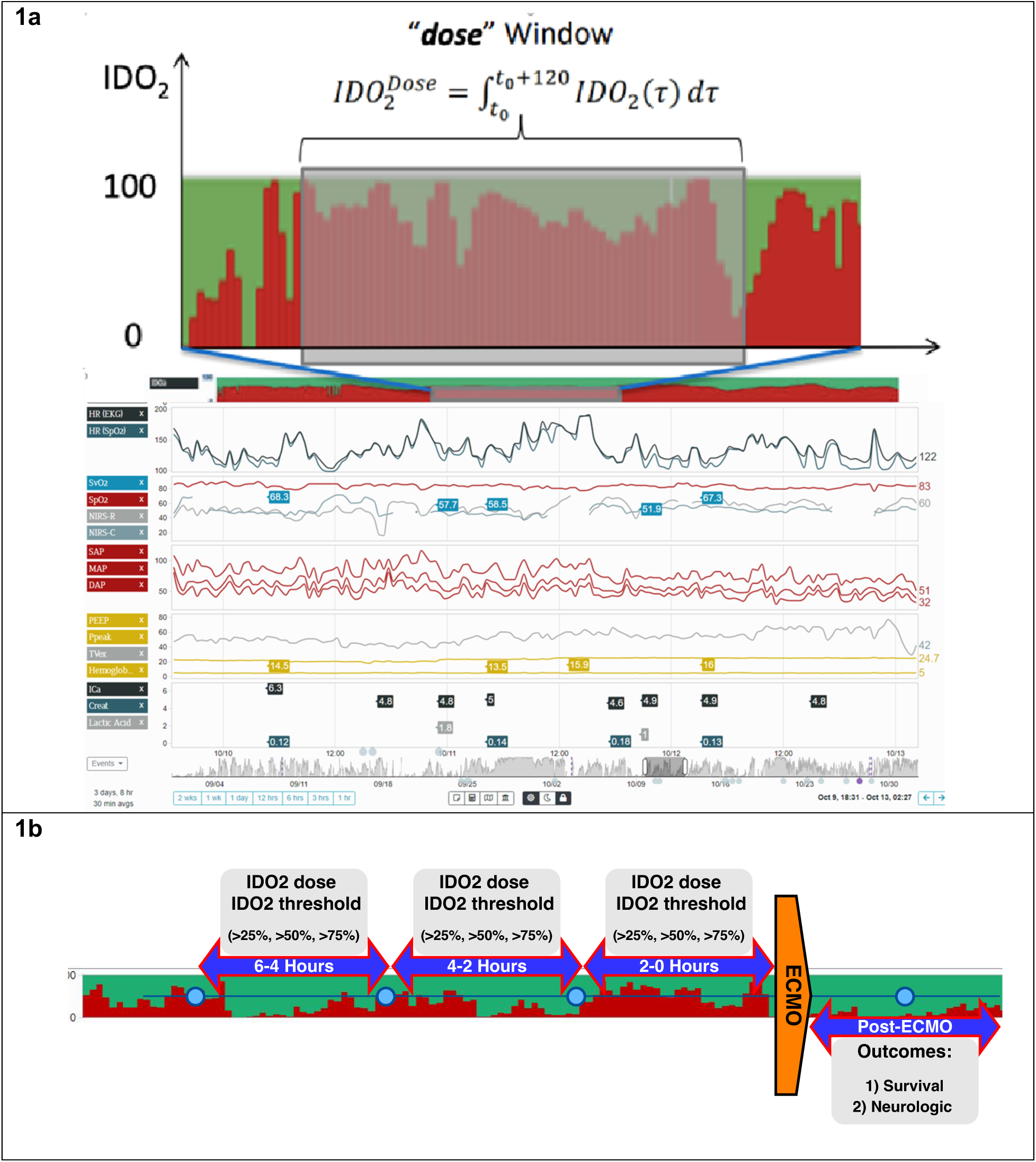
Pre-ECMO iDO2 Visualization, Trends and Associated Outcomes. **1a**. iDO2 dose and thresholds were analyzed in 2-hour windows beginning 6 hours prior to a documented cardiac arrest or ECMO deployment. Post-ECMO outcomes were evaluated in terms of survival and both short– and long-term neurologic outcomes, incorporating head imaging, EEG, and functional status scoring. **1b.** iDO2 dose and thresholds were analyzed in 2-hour windows beginning 6 hours prior to a documented cardiac arrest or ECMO deployment. Post-ECMO outcomes were evaluated in terms of survival and both short– and long-term neurologic outcomes, incorporating head imaging, EEG, and functional status scoring. ECMO, extracorporeal membrane oxygenation; EEG, electroencephalography; CT, computed tomography.

### Outcomes

Primary outcomes included short-term neurologic outcomes: 1) mortality, 2) abnormalities on electroencephalogram (EEG) while on ECMO including presence of seizures and characterization of background EEG activity, 3) head imaging abnormalities on ECMO by both head ultrasound (HUS) and computerized tomography (CT). Secondary outcomes of longer term neurologic abnormalities included: a) post ECMO magnetic resonance (MRI) head imaging abnormalities and b) functional status as graded by Functional Status Scores (FSS) at hospital discharge from review of the EHR discharge documentation (**Figure 1b**). FSS is a validated scoring tool to assess dysfunction or use of assistance devices in different functional domains like communication, respiratory, and motor[7] and gives a score range of normal (6) to varying degrees of abnormal (30 being worst). In this study, FSS was considered abnormal if it had worsened by discharge compared to admission, defined as a score in the upper 50th percentile relative to the admission median score.

EEG data were collected from available EEG reports, as clinically indicated, as during the study period EEG monitoring for all ECMO patients was not yet our institutional standard of practice. EEG studies were performed using Nihon Kohden digital EEG systems (Nihon Kohden, Tokyo, Japan) with an extended international 10–20 electrode placement with minor modifications when necessary due to head positioning. The EEG reports provided information regarding duration of EEG monitoring, background elements, seizure presence, and time of seizure onset. Along with the EEG waveforms, these reports were independently reviewed by two study investigators (RG, SBH), and then categorized into favorable and unfavorable results based on a previously reported and validated prediction tool[8–13].

Similarly, two study investigators (RG, SBH) assigned an injury severity score (normal, mild, moderate, or severe) for each patient based upon collective clinical imaging data (e.g., head ultrasound, head CT, brain MRI). The mild designation was assigned in cases with low volume of injury (e.g., punctate hemorrhages), unrelated structural findings designated by the radiologist (hypomyelination, delayed sulcation, ventriculomegaly, etc.), or likely normal variants. Moderate or severe designations involved larger injury volume, injuries requiring neurosurgical intervention (thrombosis or progressive ventriculomegaly), and two or more mild category injuries[8–13]. To account for the anticipated high incidence mortality when assessing functional outcomes, we developed a composite outcome of both FSS and survival. Survival with an FSS below the median was considered a good functional outcomes and non-survival or an FSS above the median was considered a poor outcome.

For both EEG and head imaging, one investigator initially reviewed reports with scores independently confirmed by second investigator. Disagreement regarding injury severity was reconciled by review of data by both investigators together for consensus. MRI or CT findings superseded the imaging grade of earlier HUS studies if both study types were available.

### Statistical Analysis

The relationship between these primary outcomes and iDO2thresholds and dose were statistically analyzed using SAS® 9.4 version (SAS Institute Inc., Cary, NC, USA). Qualitative variables were presented as n (%) and quantitative variables were presented as median with interquartile range (IQR) and range [min – max] or mean and standard deviation (SD) when necessary to best display the study findings. Univariate logistic regression was used to determine the relationship between IDO2 dose or threshold with mortality, head imaging, EEG and FSS. Significant association was calculated with likelihood ratio chi-square test of univariate logistic regression and significant odds ratio with profile-likelihood confidence intervals. P<0.05 was considered as significant.

## RESULTS

### Patient Characteristics and IDO2 variables

From January 1, 2013 to September 30, 2017, 219 patients underwent ECMO of which 56 met study inclusion criteria. Insufficient data to calculate IDO2 values led to exclusion of 9 additional patients, leaving a study cohort of 47 patients (**Figure 2**). The study cohort was 60% female with median age and weight at ECMO initiation of 16 days [IQR 6-112] and 3.3kg [2.8-4.8], respectively. Median ECMO duration was 135 hours [IQR 64-262] with 43 (91%) supported on veno-arterial (VA) ECMO and the remaining 4 (9%) on veno-venous (VV). The leading indication for ECMO was cardiac failure in 29 (62%) patients, with the remaining being respiratory failure 10 (21%) and pulmonary hypertension 8 (17%). ECMO following cardiac arrest, otherwise known as extracorporeal cardiopulmonary resuscitation (ECPR), comprised 16 patients (34%) while the remaining 31 (66%) underwent elective ECMO initiation. The overall mortality included 26 (55%) patients (**Table 1).** There was a higher proportion of congenital heart disease in the non-survivors 20 (77%) compared to the survivors 9 (42%), p=0.03. Additionally, there was a higher incidence of pre-ECMO cardiac arrest and ECPR in the non-survivors, 14 (53%) and 13 (50%) compared to the survivors 3 (14%) and 3 (14%), p= 0.006 and 0.014 respectively.

**Figure 2:**
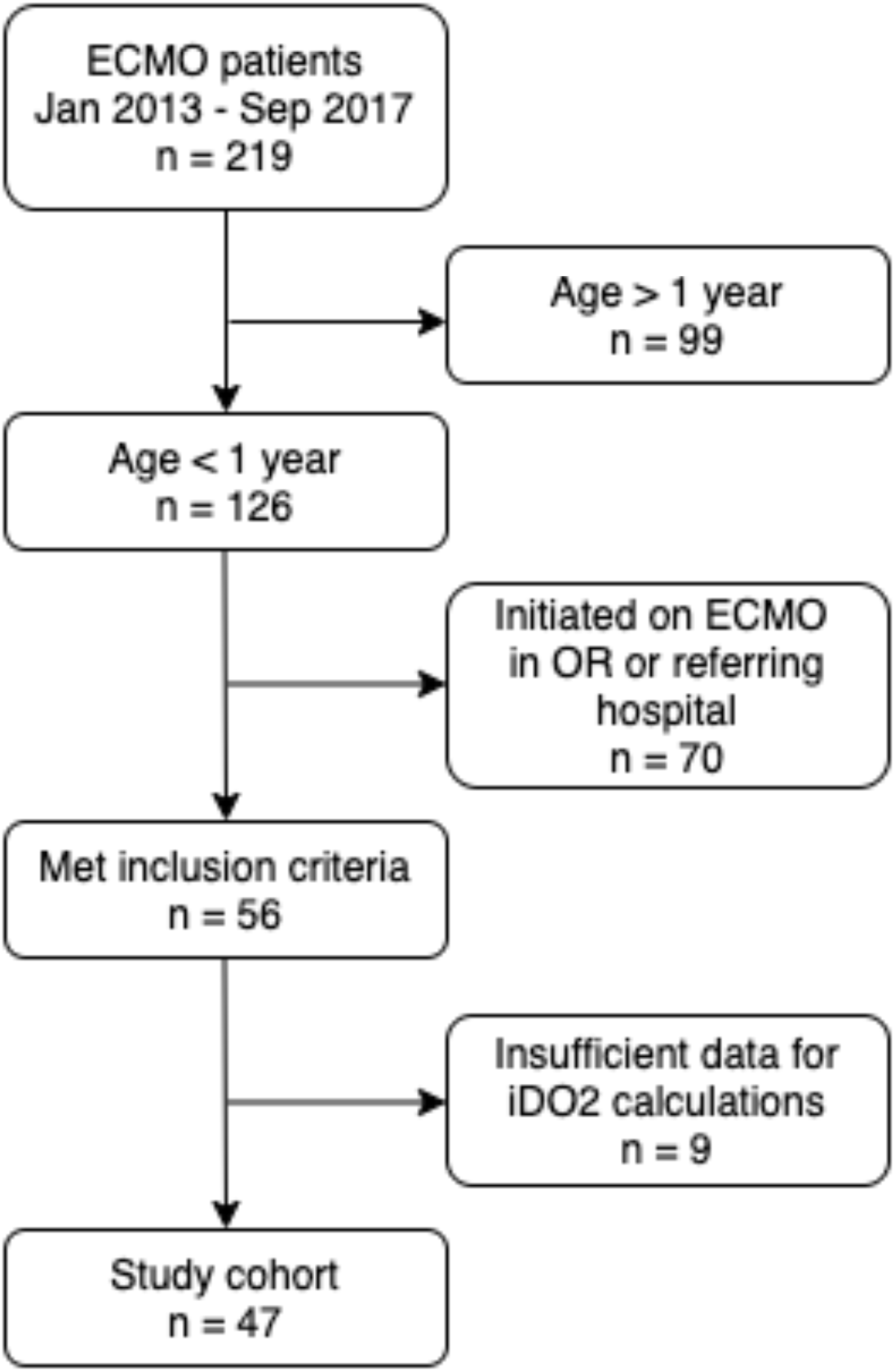
Patient Cohort Selection: CONSORT Diagram. A total of 219 patients were identified from 2013 to 2017, of which 126 were infants. Of these, 46 infants met the inclusion criteria and had sufficient data available to generate iDO2 calculations for analysis.

**Table 1:**
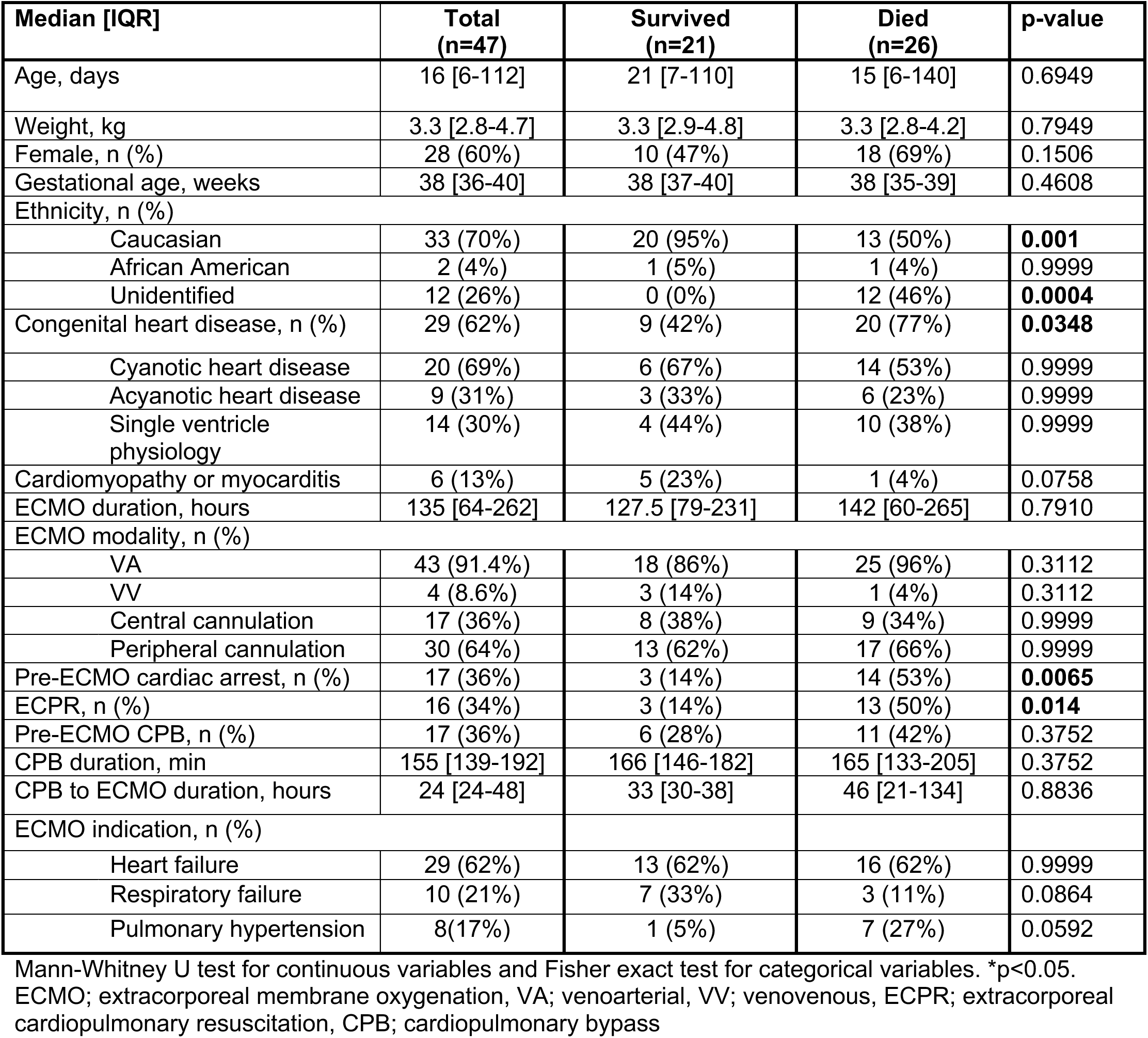
patient demographics and IDO2 variables.

The mean time spent above iDO2 probability thresholds of 25%, 50% and 75% increased in the time intervals closest to ECMO initiation. The duration of time spent above the iDO2 threshold of 25% was mean±SD 18±32, 19±32, and 56±158 minutes, at the 6-4 hours, 4-2 hours, and 2-0 hours prior to ECMO initiation, respectively, p = 0.149 (**Figure 3A**). For the time above the threshold of 50%, the durations were mean±SD 9.4±21, 7.8±17.88, and 23.4±31.4 minutes, at the 6-4 hours, 4-2 hours, and 2-0 hours prior to ECMO initiation, respectively, p = 0.0003 (**Figure 3B**). Lastly, the times spent above the iDO2 threshold of 75% were mean±SD 2.7±8.3, 2.2±7.4, and 16.3±25.3 minutes, at the 6-4 hours, 4-2 hours, and 2-0 hours prior to ECMO initiation, respectively, p<0.0001 (**Figure 3C**). There was a statistically significant increase in the iDO2 dose in the time intervals closer to ECMO initiation, mean±SD 12.23±18.12 in the 6-4 hours, 19.85±14.78 in the 4-2 hours and 31.63±31.38 in the 2-0 hours prior to ECMO initiation, p = 0.003 (**Figure 3D**).

**Figure 3:**
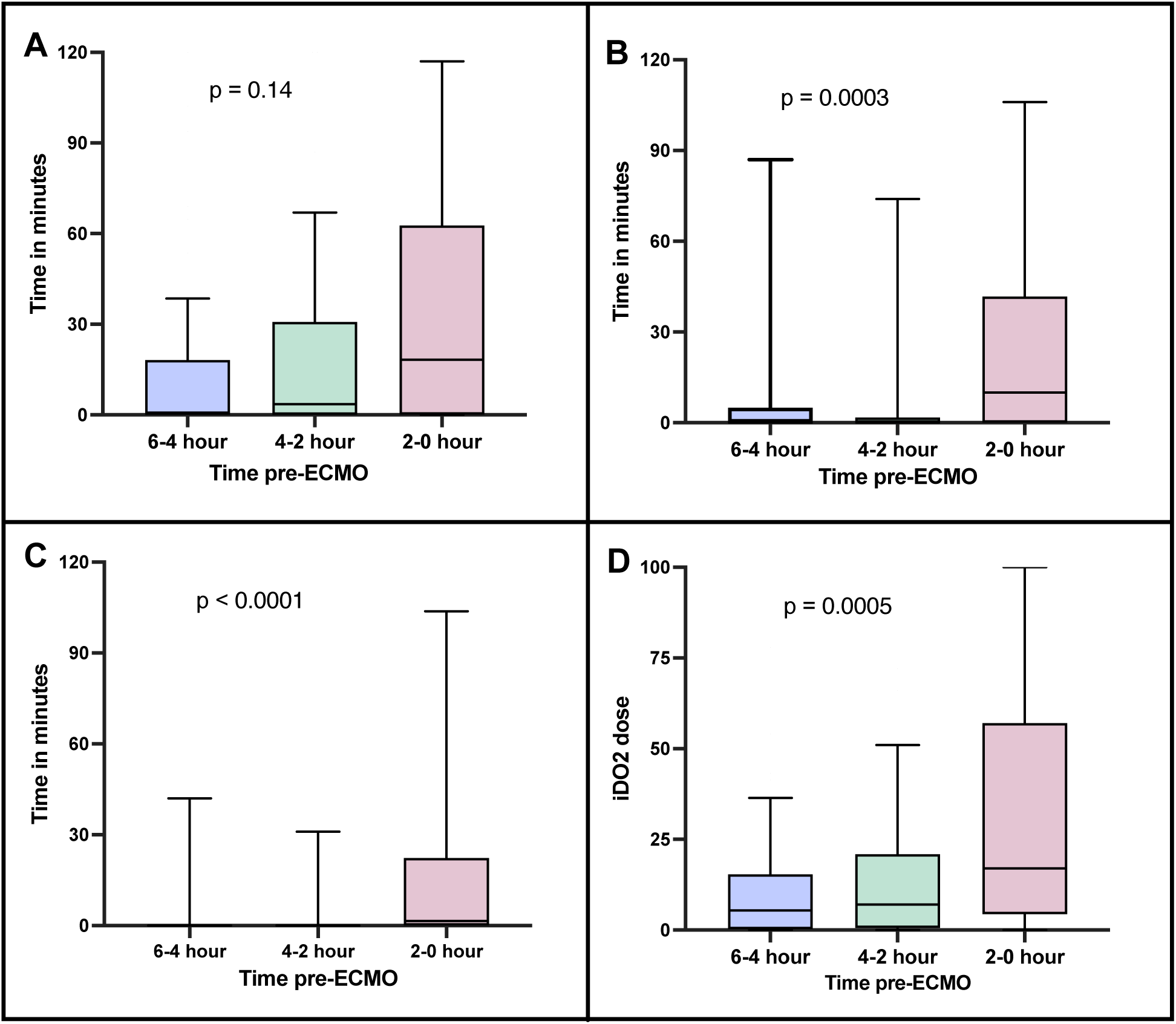
Trends in Pre-ECMO iDO2 Dose and Time Spent Above Thresholds. **3a**. The iDO2 dose showed a progressive increase as patients neared ECMO cannulation. **3b-d.** As iDO2 dose increased, there was a corresponding rise in the duration of time spent above the studied iDO2 thresholds. ECMO, extracorporeal membrane oxygenation.

### Outcomes

On-ECMO EEG data was available in 32 patients (68%). Seizures were diagnosed in 15 (47%) of these patients and background asymmetry in 17 (53%) (**Table 2**). Of note, prior to ICU admission, 6 of the 47 (13%) patients included in the study had been maintained antiepileptic medications prior to ECMO, suggesting pre-existing epileptiform conditions. On ECMO HUS was available for all patients and abnormalities were found in 33 (70%) patients. Head CT was available in 18 (38%) cases and abnormalities were found in 12 (67%) patients (**Table 2**). The most common abnormality was hemorrhage in 8 (67%) patients, followed by ischemia in the remaining 4 (33%) cases. Associated diffuse cerebral edema was present in 2 (17%) cases. Pre-ECMO CT was available in 8% of cases of which a single patient had a noted abnormality (chronic signs of encephalomalacia). All patients had HUS imaging on ECMO with 33 (70%) with recorded abnormalities.

**Table 2.** – patient outcomes:

| Outcome measure, n (%) | Total<br>(n=47) | Survivors<br>(n=21) | Non-survivors<br>(n=26) | p-value |
| --- | --- | --- | --- | --- |
| **On ECMO EEG, n=32 (68%)** |  |  |  |  |
| Seizures | 15 (47%) | 4 (19%) | 11(42%) | 0.1208 |
| EEG asymmetry | 17 (53%) | 5(24%) | 12(46%) | 0.1376 |
| **On ECMO head CT, n=18 (38%)** |  |  |  |  |
| Head CT abnormalities | 12 (67%) | 1(5%) | 11(42%) | **0.0057** |
| **On ECMO head US, n=47 (100%)** |  |  |  |  |
| Head US abnormalities | 33 (70%) | 14(67%) | 19 (73%) | 0.5274 |
| **Post-ECMO head MRI, n=20 (43%)** |  |  |  |  |
| Head MR abnormalities | 15 (75%) | 15 (75%) | NA | NA |
| **FSS at hospital discharge, median [IQR], n=21 (46%)** | 9 [8, 11] | 9 [8, 11] | NA | NA |
| Fisher exact test for categorical variables. *p<0.05.<br>ECMO; extracorporeal membrane oxygenation, EEG; electroencephalogram, CT; computer tomography, US; ultrasound, MRI; magnetic resonance imaging, FSS; functional status score |  |  |  |  |

Of note, there were differences in the EEG or head imaging outcomes between survivors and non-survivors except for head CT findings where non-survivors had significantly higher incidence of abnormalities, 11 (42%), compared to survivors, 1 (5%), p=0.005. Following ECMO, MRI imaging was available in 43% of cases, 75% of survivors. Abnormalities were noted in 75% of cases, including hemorrhage (40%), FLAIR hyperintensities (13%), and diffusion restriction, sinus venous thrombosis, or encephalomalacia in 7% each.

Baseline FSS was available on 33 (70%) patients and the median was found to be 10 [IQR 9-14] (**Table 2**). Of survivors, FSS at hospital discharge was available in 21 (100%) of patients. The median FSS at discharge was 9 [IQR 8-11]. Of survivors, 75% of patients were discharged with new support technology (all accounted for as part of FSS[7]). This was most commonly a nasogastric tube (53%) gastric or gastric-jejunal tube (26%), and tracheostomy in 16% of survivors, of whom 5% also required mechanical ventilation at discharge.

### Associations of iDO2 with various outcomes

When assessing the relationship of the various iDO2 variables and mortality, patients who did not survive their ECMO course had significantly higher iDO2 dose in the 2 hours immediately prior to ECMO initiation (Mean±SD 40.17±34.8 vs 18.46±21.18, p=0.02) and more time spent above an iDO2 threshold of 75% (Mean±SD 23.6±28.2 vs 5.61±15.58 minutes, p=0.01), **Figure 4A**. When analyzing the relationship of iDO2 variables and short term neurologic outcomes, patients with abnormalities on head imaging data had significantly higher iDO2 in the 6-4 hours interval prior to ECMO initiation compared to the patients with normal head imaging (Mean±SD 16.29±15.6 vs 1.22±2.72, p=0.04), **Figure 4B**. For the EEG outcomes, there was patients with EEG asymmetry had significantly higher iDO2 dose, time spent above an iDO2 threshold of 25%, and time spent above a threshold of 50% in the 6-4 hours interval prior to ECMO initiation (Mean±SD 15.19±19.94 vs 3.33±5.005, p=0.008, 23.34±35.90 vs 1.444±3.439 minutes, p=0.004, and 12.47±23.67 vs 0.2111±0.6333 minutes, p=0.01, respectively), **Figure 4C**.

**Figure 4:**
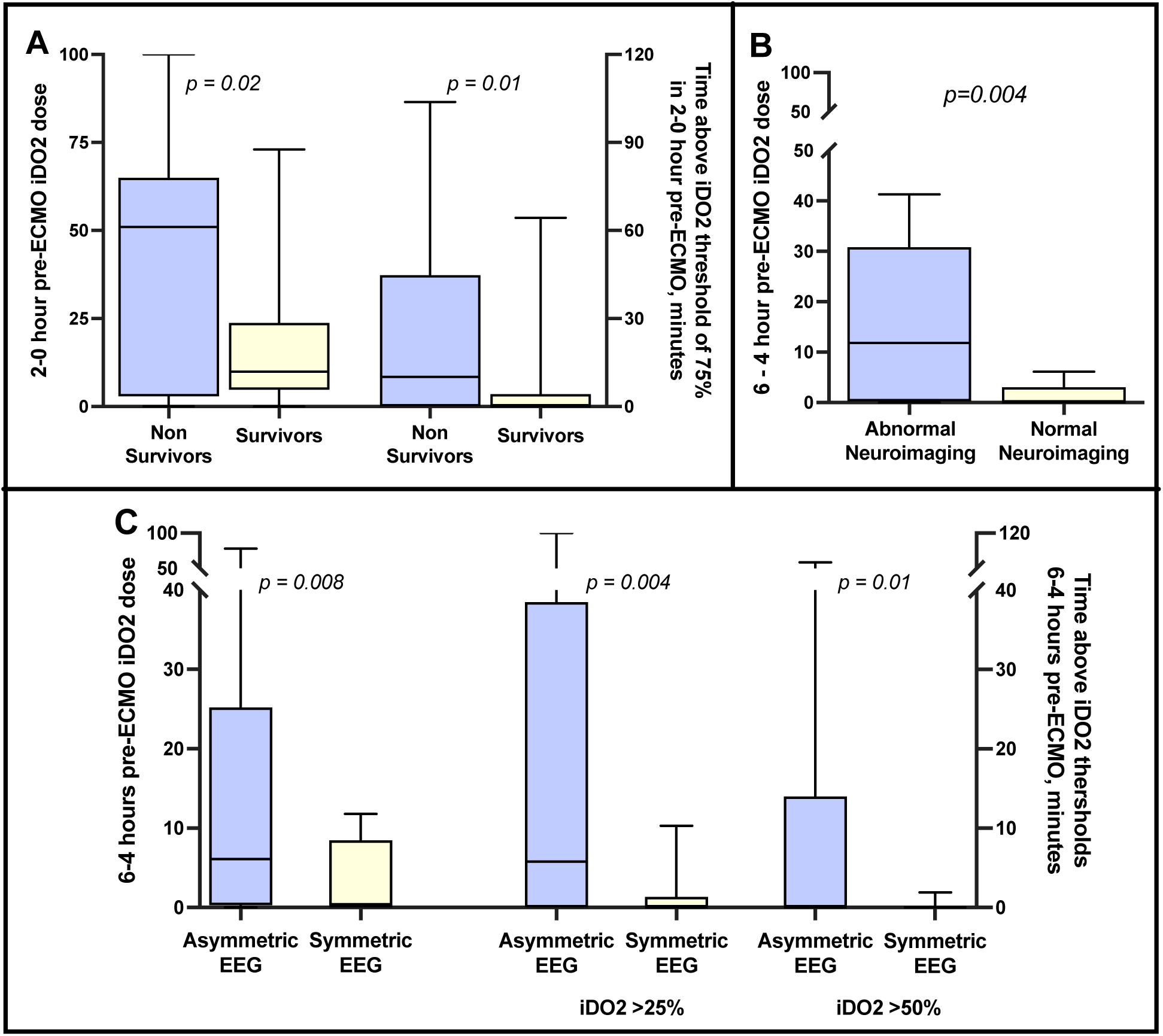
Pre-ECMO iDO2 Variables and Their Association with Neurologic Outcomes. **4a**. Analysis of iDO2 and EEG data showed that as early as 6 hours before ECMO initiation, patients with asymmetric hemisphere activity on EEG during ECMO exhibited a significantly higher iDO2 dose. Increased time spent above the 25% and 50% iDO2 thresholds before ECMO was associated with a greater likelihood of asymmetric EEG activity. **4b.** A heavier iDO2 dose 4–6 hours before ECMO initiation correlated with abnormal head CT findings during ECMO. **4c.** In the final 2 hours before ECMO cannulation, patients who did not survive exhibited a significantly higher iDO2 dose. Increased mortality was associated with greater time spent above the 75% iDO2 threshold. ECMO, extracorporeal membrane oxygenation; EEG, electroencephalography; CT, computed tomography.

Given that the median FSS at discharge 9, a cutoff for FSS <10 for survivors was used to identify a good functional outcome and death or: =10 was used to define a poor functional outcome. A mixed effects model for the change in iDO2 dose over the 3 examined timepoints for the good vs poor functional outcome was significant (p<0.001), (**Figure 5**). Evaluating the feature combinations most predictive of with composite functional outcome, a logistic regression model including the time spent above the 3 iDO2 thresholds of 25%, 50% and 75% at the various studied time intervals of 6-4 hours, 4-2 hours and 2-0 hours prior ECMO initiation had the best predictive performance with an area under the receiver operator curve (AUROC) of 0.81, 95% confidence interval 0.7-0.92 and p value of 0.0002 (**Figure 6**).

**Figure 5:**
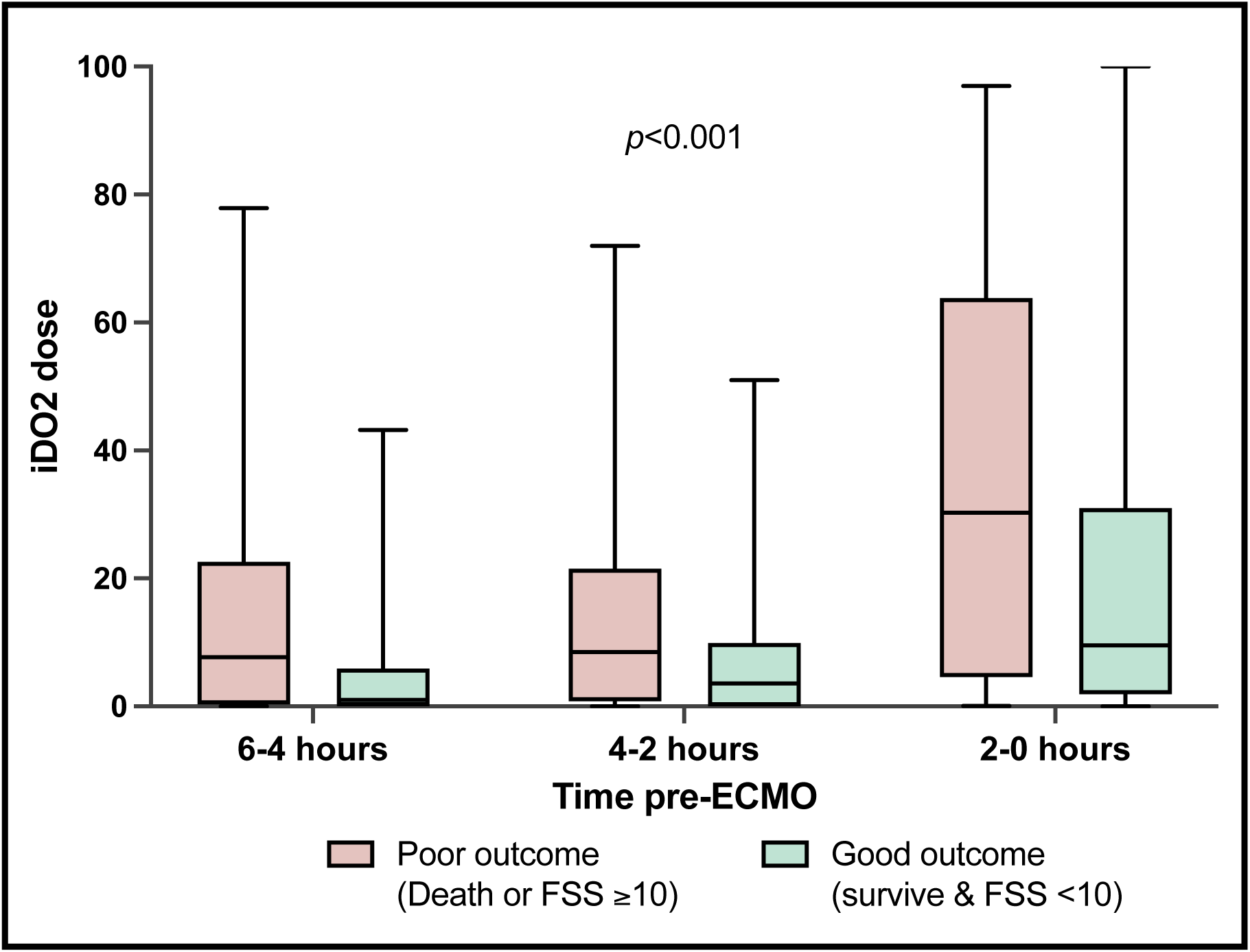
Pre-ECMO iDO2 and Functional Status Score (FSS) or Mortality. Starting as early as 6 hours prior to ECMO cannulation, patients with poor outcomes (abnormal FSS or death) demonstrated a significantly higher and progressively increasing iDO2 dose. ECMO, extracorporeal membrane oxygenation; FSS, Functional Status Score.

**Figure 6:**
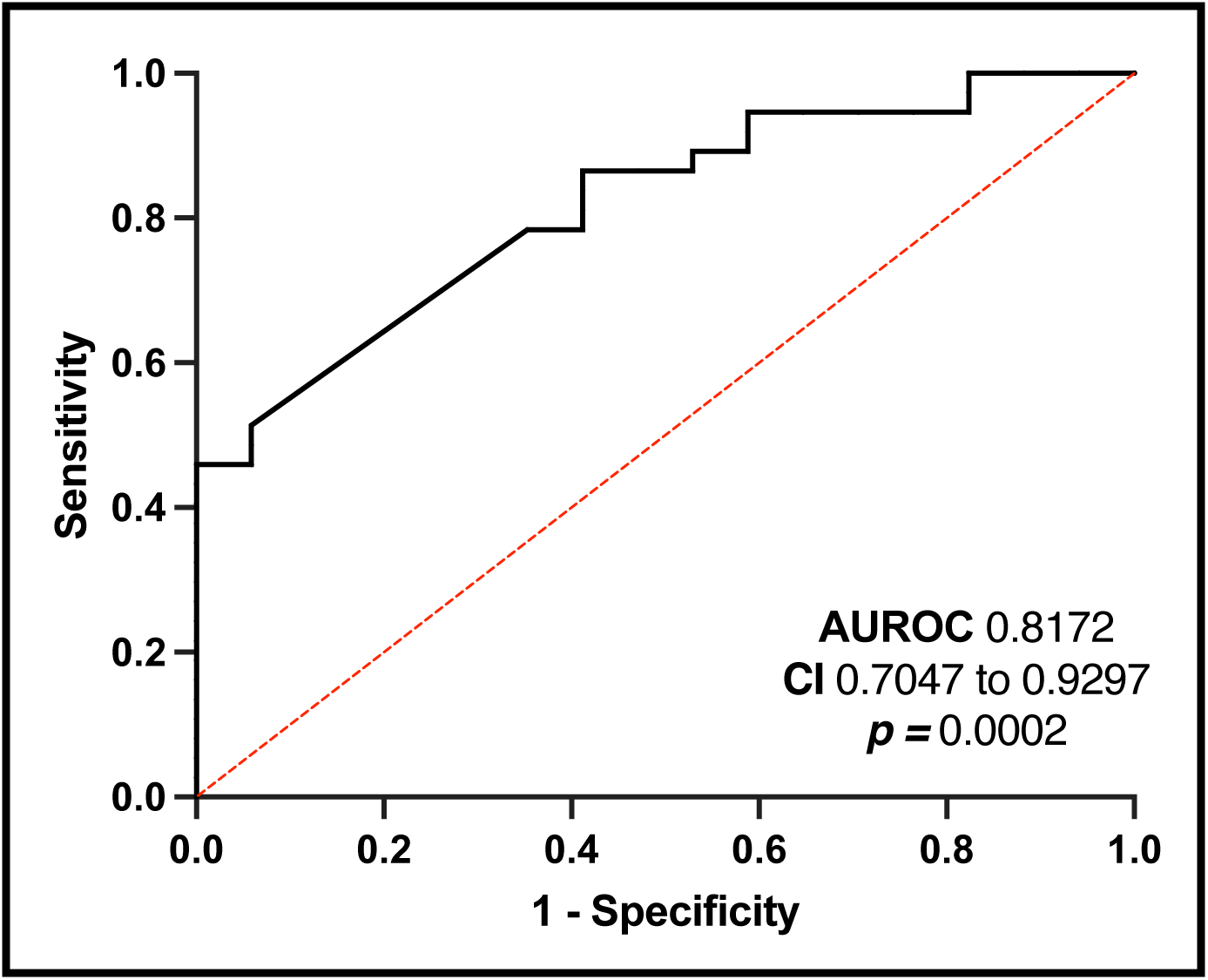
Logistic regression model of time above iDO2 thresholds to predict the composite outcome. A multivariate logistic regression model including the time spent above the various iDO2 thresholds, 25%, 50%, and 75%, at the 3 timepoints prior to ECMO initiation to discriminate for the composite outcome of survival with an FSS score above the median. The model had a strong predictive performance with an AUROC of 0.81 (95% CI: 0.7 – 0.92, p = 0.0002). ECMO, extracorporeal membrane oxygenation; FSS, Functional Status Score; AUROC, area under the receiver operator curve.

## DISCUSSION

ECMO has become an invaluable tool in the management of life threatening pediatric cardiac and or respiratory failure. Despite its increasing use, the decision to initiate ECMO has highly subjective lacking objective tools to identify the ideal timing of ECMO initiation when the benefits of ECMO outweigh and its potential risks. In this pilot study, we demonstrated that the duration and depth of inadequate oxygen delivery, up to 4 to 6 hours prior to ECMO initiation, as measured by high resolution physiologic data and iDO2 analytics, is associated with worse outcomes including death, neurological complications and worse functional outcomes at hospital discharge.

The incorporation of ECMO into the management of pediatric cardiac and respiratory failure has significantly improved patient outcomes, transforming previously fatal conditions into recoverable ones[14]. While survival remains a key goal, reducing associated morbidity and preserving quality of life are increasingly important considerations. Earlier deployment of ECMO in certain patients could enhance end-organ oxygen delivery, improving survival and reducing complications. However, determining the optimal timing for ECMO initiation remains challenging due to the lack of objective decision-support tools, a gap this study aims to address.

Clinical recognition of impaired oxygen delivery, a critical factor in ECMO timing, is difficult even for experienced clinicians[15]. Cardiac arrests in pediatric ICUs, often preceded by progressive deterioration, occur at rates of 2.6–6% and carry mortality rates exceeding 47%[16], reaching 64% under specific high-risk circumstances[17]. Existing ECMO prognostication tools, such, focus primarily on mortality and rely on static, registry-based data, neglecting dynamic patient trajectories and non-mortality outcomes like neurologic and functional impairments[18]. Pediatric ECMO survivors face significant long-term challenges, including motor impairments, cognitive delays, and neurodevelopmental deficits[19,20]. As such, existing tools also fail to provide real-time decision support, limiting their bedside utility.

Studies have repeatedly linked markers of worsening end organ function, mostly likely secondary to the severity of impaired systemic oxygen delivery and lack of physiological reserve, to worse post-ECMO outcomes[21–27]. iDO2 analytics represents a promising near real-time aid for quantifying impaired systemic oxygen delivery. By integrating high-resolution physiologic data, it provides estimates of oxygen delivery providing an additional tool in managing critically ill children. Studies have shown iDO2 correlates with key clinical indicators, such as mixed venous oxygenation, lactate levels[28], and cardiac ICU length of stay[29–31], and can predict cardiac arrest[4]. Our pilot data further support that higher iDO2 doses and durations spent above various iDO2 thresholds, even hours prior to ECMO initiation, correlate with worse outcomes. It is also important to note that patients who received ECPR in our study had a six-fold higher mortality rate compared to those electively placed on ECMO, highlighting the potential of iDO2 to guide earlier, safer ECMO deployment.

iDO2 is not a replacement for clinical judgment but a tool to enhance it, offering standardized, objective physiologic measurements to support ECMO decisions. It may alert less experienced providers to impending risks and reduce variability in ECMO initiation practices across institutions. Although iDO2 dose was not available to the healthcare team in this study, our findings suggest that its integration into clinical workflows has the potential improve outcomes and reduce morbidity by identifying the optimal time for ECMO initiation. Emerging evidence further indicates its potential to positively impact surgical and critical care outcomes, warranting further investigation into its role in real-time clinical decision-making[32].

Limitations of this study include its retrospective design and relatively small sample size. This makes it challenging to fully assess the clinical context in which practitioners made decisions and how these influenced patient outcomes and ECMO timing. The modest cohort size also limits the ability to analyze every pre-ECMO time point and outcome comprehensively. The study’s lack of a control group is another limitation, as patients were selected from an institutional registry of infants who had already received ECMO, preventing comparisons with a matched non-ECMO cohort with similar iDO2 variables. Additionally, potential selection bias may have influenced analyses of on-ECMO EEG and head CT data, as these tests were likely ordered for patients who were clinically deemed to require them. Larger, multi-institutional, prospective trials are needed to validate these pilot findings and evaluate their applicability to more heterogeneous ECMO cohorts.

Furthermore, additional studies to analyze the differences in subsets of ECMO populations will help specify which patient cohort would most benefit from early detection of low oxygen state and early ECMO deployment. These for example can include neonates, children, adolescents, those requiring venovenous or venoarterial ECMO as well as a range of pathologic etiologies such as sepsis, acute respiratory distress syndrome or immunocompromised children.

## CONCLUSION

In conclusion, in this study we show an association between the duration and severity of impaired oxygen delivery, hours prior to ECMO initiation, as quantified by iDO2 metrics with worse mortality and neurological outcomes. These findings lay the groundwork for future multi-center studies to validate our findings on larger, more heterogenous ECMO populations. The incorporation of metrics of impaired oxygen delivery into objective decision support tools to guide the timing of ECMO initiation before the development of severe end organ damage, has the potential to improve both short and long-term outcomes of pediatric ECMO.

## Data Availability

All data produced in the present study are available upon reasonable request to the authors

## Acknowledgements

We would like to thank the Etiometry team (Etiometry Inc., Boston MA) for their insight and support in the data analysis and management.

